# Data dashboard to support meta-analyses on influenza vaccine effectiveness across regions, influenza seasons and outcomes

**DOI:** 10.1101/2025.06.12.25329122

**Authors:** David Hendrickx, Keira Lee Rice, Hunter Baggen, Jacopo Frallicciardi, Joana Carvalho Moreira de Mello, Boris Julg, Anita H.J. van den Biggelaar, Jaap Goudsmit, Anna L. Beukenhorst

## Abstract

Evaluating influenza vaccine effectiveness is essential for identifying gaps in influenza prevention strategies.

We developed an open-source dashboard-ready dataset with vaccine effectiveness data from 45 index countries in the inter-pandemic period between 2011 and 2019, based on a systematic literature review. We show how it can be used to estimate pooled vaccine effectiveness using fixed and/or random-effects meta-analyses.

The full dataset contains 9,632 vaccine effectiveness estimates from 239 peer-reviewed studies. Individual data points are available on vaccine valency, type, and dose. The meta-analyses dataset can be subset by model, geographic region, influenza season, measure (adjusted, unadjusted), outcome (lab-confirmed infection, influenza-like illness, influenza-associated complications) age groups, comorbidity- or age-based risk groups, influenza type, and medical attendance type.

As an example, we used a subset (with data from 2,128,431 participants) of this data to calculate the fixed effects pooled vaccine effectiveness for laboratory-confirmed influenza, which was 39.4% (for all). Vaccine effectiveness was lowest in the elderly (65+: 27.4%) compared to the general population; against A/H3N2 (23.9%) compared to A/H1N1 (49.0%) or B (46.7%); and in 2014, when there was mismatch between the circulating A/H3N2 strain and vaccine antigen.

Our open-source datasets enable policy makers and researchers to assess vaccine effectiveness in different regions and contexts. Given the notably low vaccine effectiveness for the elderly, complementary strategies to further reduce the yearly worldwide burden of influenza infections are needed.

## Introduction

Every year, seasonal influenza affects approximately one billion people, leading to 3–5 million severe cases of illness, and 290,000 to 650,000 deaths. The primary method of prevention is through seasonal influenza vaccines, regularly updated to match predicted circulating viral strains^1^. Vaccines lessen the impact of influenza, yet do not necessarily prevent infection^2^ or the economic consequences of reduced productivity and increased healthcare in the working population^3–5^.

Vaccine effectiveness of the seasonal influenza vaccine varies. Vaccines tend to be less effective for populations with the highest burden of severe influenza; the elderly^6,7^ and those who are immunocompromised^8,9^. The majority of influenza-related hospitalizations (50-70%) and deaths (70-85%) occur in these population groups^10^. Vaccine effectiveness further depends on the outcome: vaccines are more effective against mortality than morbidity^11^.

Individual studies on vaccine effectiveness therefore yield a broad spectrum of vaccine effectiveness estimates. In the United States of America, seasonal influenza effectiveness reportedly ranges from 19% to 60%^12^. Part of the variability reflects a true difference in vaccine effectiveness according to the abovementioned factors. Another part is due to the skewing of vaccine effectiveness estimates due to study-specific factors such as choice of endpoint, population size and demographics season.

Given the critical role of influenza vaccination in public health strategies, it is essential to (1) use the highest level of epidemiological evidence on vaccine effectiveness – the systematic literature review with meta-analysis, and (2) pool scientific efforts in the form of open-source data to allow different stakeholders to analyse vaccine effectiveness for different purposes and synthesize the available evidence methodically. Identifying drivers of low vaccine effectiveness can then help shape new interventions and targeted public health strategies to reduce the burden of influenza.

In this study, we performed a systematic review of the literature on the effectiveness of seasonal influenza vaccines. We present a publicly available dataset with vaccine effectiveness estimates over nine consecutive seasons. We showcase its usage with meta-analyses on a subset of studies containing a total of 2,128,431 study participants, calculating pooled vaccine effectiveness in various demographic groups and influenza seasons, and for different influenza subtypes.

## Methods

We conducted a systematic literature search for studies that report adjusted vaccine effectiveness estimates and their 95% confidence intervals for laboratory-confirmed influenza and/or influenza-like illness. Laboratory-confirmed influenza was defined as a patient who presented with influenza symptoms and was found to be influenza-positive by Polymerase Chain Reaction/Nucleic Acid Amplification Testing (PCR/NAT), virus culture, immunofluorescence test, or antibody assessment in paired sera. We defined influenza-like illness as patients who presented with clinical signs and symptoms, but where no laboratory confirmation other than a Rapid Antigen Test (RAT) was conducted. Studies that reported only relative vaccine effectiveness, immunological outcomes, or unlicensed vaccines were excluded. Full inclusion and exclusion criteria are provided in the methodological supplement (see Appendix I).

We used the web-based tool Rayyan^13^ for study screening and selection, selecting relevant studies on title and abstract first and the remainder on full text.

### Search strategy

We used the following search terms in the Medline bibliographic database: (Flu[tiab] OR “Influenza-like illness”[tiab] OR “Influenza A virus”[Mesh] OR “Influenza B virus”[Mesh] OR “Influenza, Human”[Mesh]) AND (“Vaccine Efficacy”[tiab] OR “Vaccine Effectiveness”[tiab] OR “Vaccine Efficacy”[Mesh]). We applied the following limits: 2012/01/01:2022/12/31[Date - Publication] AND (“humans”[MeSH Terms] AND (“dutch”[Language] OR “english”[Language] OR “french”[Language] OR “german”[Language] OR “spanish”[Language])) NOT “review”[Publication Type].

### Data extraction

We extracted all reported adjusted and unadjusted vaccine effectiveness estimates, with confidence intervals, from 2011/12 (northern hemisphere) and 2012 (southern hemisphere) onwards, both for the general population and for risk groups. Vaccine effectiveness was defined as the relative reduction in laboratory-confirmed influenza or influenza-like illness. Influenza vaccine effectiveness was derived from reported odds-ratios (ORs) and expressed as a percentage using the formula (1-OR)*100. This percentage represents the proportionate reduction in risk between vaccinated and unvaccinated groups for either of these two outcomes.

We extracted study characteristics as well as information on data sources used, study population, vaccine, and statistical methodology. These characteristics included the influenza virus type, subtype or lineage the estimate was relevant to, and the adjustments that were applied in the case of adjusted vaccine effectiveness estimates that were extracted. We used Covidence (Covidence systematic review software, Veritas Health Innovation, Melbourne, Australia. Available at www.covidence.org) and MS Excel (Microsoft Corporation. Available at www.microsoft.com/excel) for data extraction. Data extraction was independently conducted and reviewed by two researchers, and checks for missing values and data entry errors (e.g. vaccine effectiveness above 100%) were conducted in R.

### Meta-analysis

We conducted meta-analyses using the estimates of effect and their 95% confidence intervals extracted directly from the included test-negative design studies only, as per Cochrane meta-analysis guidelines for data from non-randomised studies^14^. This enabled us to perform meta-analyses on reported adjusted vaccine effectiveness estimates rather than crude estimates only, as well as allowing us to include vaccine effectiveness estimates for which summary data was not provided (see Appendix I).

The unit of analysis was vaccine effectiveness for a given country and influenza season combination. Aggregated vaccine effectiveness estimates for a combination of countries or influenza seasons were therefore excluded from the meta-analysis, as these could be biased to seasons with stronger mismatch or different study populations. Both fixed and random (DerSimonian-Laird) effects meta-analysis models were run, weighing individual study results by the inverse of their variances.

A test of heterogeneity was performed using a chi-square test at a significance level of p<0.05. These were reported with their respective I2 statistics, using 25%, 50%, and 75% cut-offs to signify low, moderate, and high heterogeneity, respectively. All meta-analyses were conducted in R, using the ‘Metafor’ package^15^.

#### Stratified meta-analyses

We performed a stratified meta-analysis by age group, by influenza season, and by influenza subtype. In addition, we performed a stratified meta-analysis for vaccine effectiveness against lab-confirmed influenza in different populations (by medical attendance type: study population consisting of outpatients presenting at a medical facility versus inpatients admitted to a hospital or intensive care unit).

The meta-analysis by age group considered studies done in populations under 5, between 5-65, and over 65 years of age. Where studies reported vaccine effectiveness estimates for narrower age bands, these were assigned to the appropriate age band, ensuring that each population was only included in one vaccine estimate to avoid data redundancy.

For the meta-analysis comparing influenza A and influenza B, vaccine estimates were grouped in ‘influenza A’ (A/H1N1 and A/H3N2) and influenza B (B/Yamagata and B/Victoria). To avoid data redundancy, not all estimates were used, especially for studies that reported multiple vaccine effectiveness estimates based on the same cases (e.g. for any influenza, influenza A, A/H1N1 and A/H3N2 separately). If studies reported estimates for all influenza A, and for each subtype separately, only the two separate subtype estimates were used. If studies reported estimates for all influenza A, and only one subtype, the estimate for ‘all influenza A’ was used. Full R-syntax for the logic behind the analysis per influenza type is provided in appendix II.

We conducted one meta-analysis by year for all studies from both hemispheres included. In addition to our main meta-analysis across all seasons and countries, we performed a second meta-analysis for studies in the United States of America only. This analysis was stratified by influenza season to enable comparison of pooled seasonal vaccine effectiveness estimates alongside available CDC-reported seasonal influenza vaccine matching information.

Figures were prepared in R version 4.2.1 and imported into Adobe Illustrator for further aesthetic adjustments.

## Results

Of 1,164 unique records that were identified, 706 were excluded following title and abstract screening and a further 219 during full-text review (reasons for exclusion in Fig. 1). Data was extracted from 239 articles: a total of 9,632 vaccine effectiveness estimates. All extracted data is publicly available and can be loaded into a shiny application for dashboard functionality (Figure 6).

**Figure 1.**
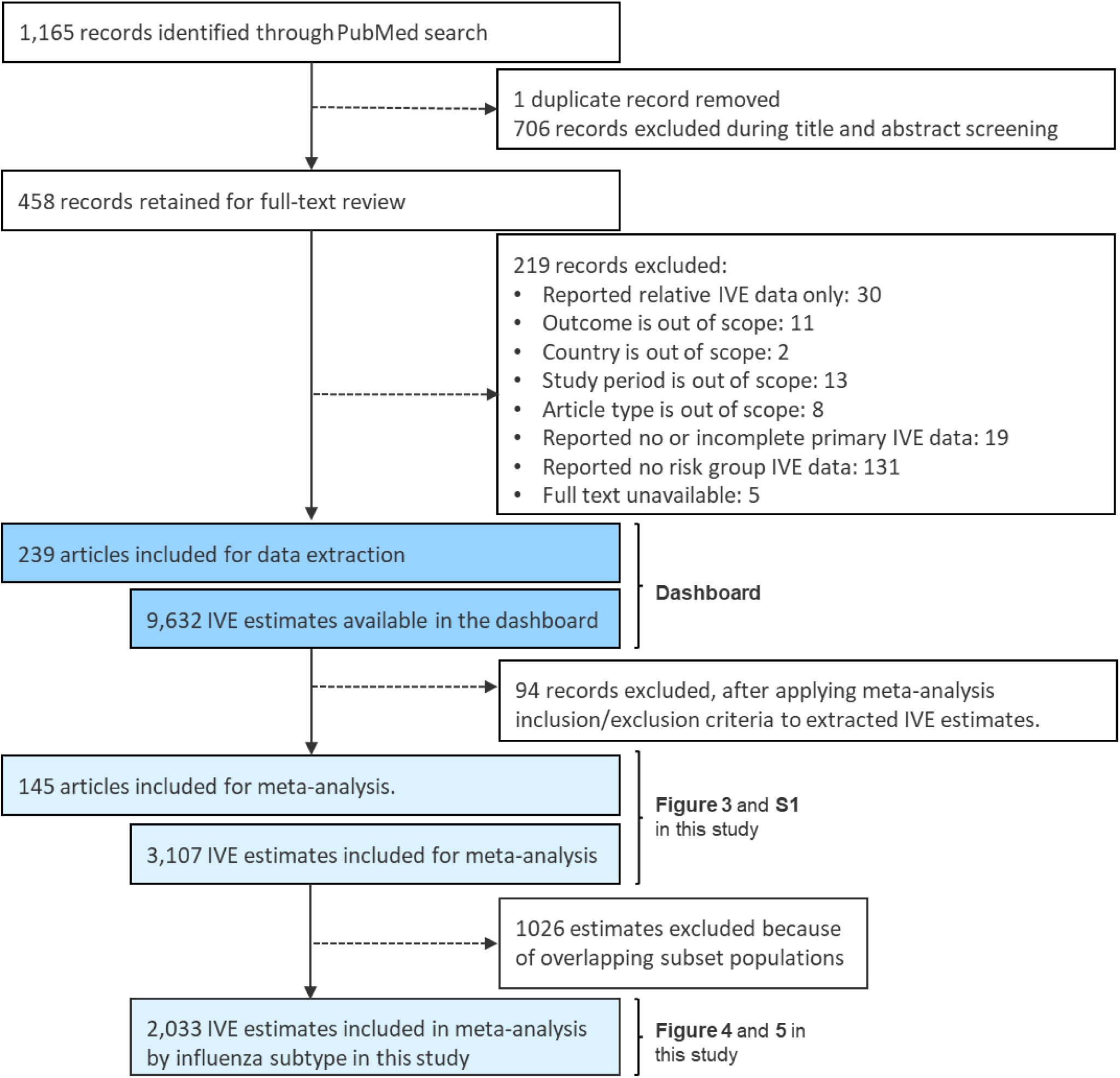
PRISMA flowchart of the systematic review process. See Appendix I for more details. IVE: influenza vaccine effectiveness.

### Study characteristics

The remainder of this article focuses on the meta-analysis of vaccine effectiveness we conducted using a subset of the extracted data and illustrates the relevance of the dataset for open data and open methods in epidemiology. We selected the 145 studies using a test-negative design, and which reported vaccine effectiveness estimates for single influenza seasons and individual countries separately (selection criteria in Appendix I). Most studies were conducted in a single country (139/145 studies; 96%); only 6 studies investigated populations in multiple countries (Fig. 2A). Most studies were conducted in the Northern Hemisphere (North America: 23%; Europe: 33%; Fig. 2A and 2C). Study size ranged between 226 and 1,250,000 participants (median = 2,421, IQR = 4,351; N_total_ = 2,128,431 participants). For the meta-analyses stratified by age group and influenza subtype, only the relevant subset of studies was used (Fig. 2C and 2D).

**Figure 2.**
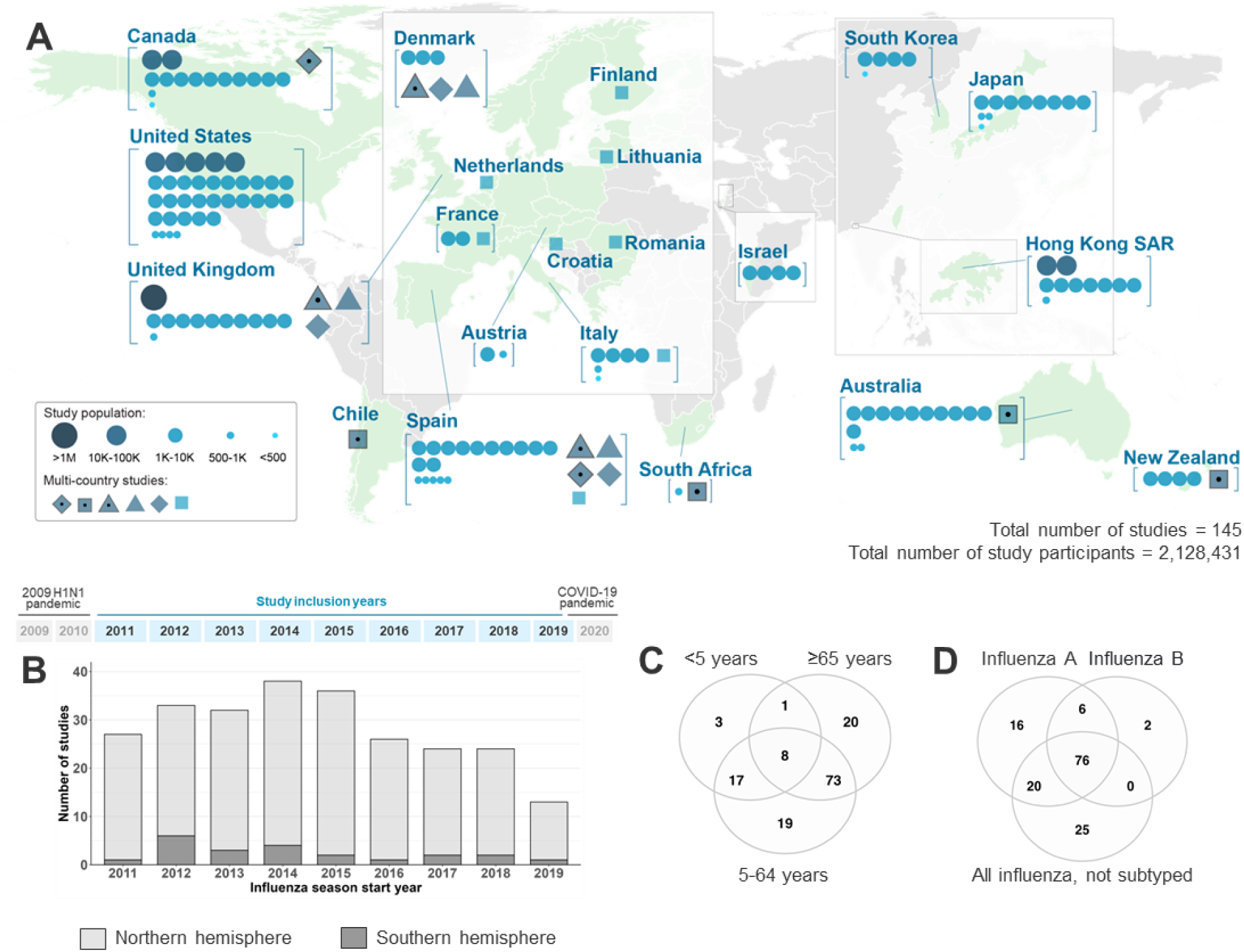
Overview of data extracted for the vaccine effectiveness meta-analysis. (A) 45 countries (green) were included in the systematic search, and 145 studies were included in the meta-analysis (each blue symbol is one study) . Circles are studies conducted in a single country; other symbols denote a unique study conducted in multiple countries. Symbol size represents study size (one study in Spain was omitted as study size was not reported). Some regions were magnified to improve the clarity of labels; Antarctica is not shown. (B) Number of studies per influenza season included in the meta-analysis, from the Northern Hemisphere (light grey) and Southern hemisphere (dark grey), respectively, by influenza season start year (*e.g*., the 2011/12 Northern Hemisphere season and 2011 Southern Hemisphere season were grouped under the 2011 influenza season start year). (C) Number of studies included in stratified meta-analyses by age. (D) Number of studies included in meta-analyses by influenza subtype.

All adjusted vaccine effectiveness estimates for lab-confirmed infection and influenza-like illness, and the relative distribution of estimates across age and influenza subtype are shown in Figure 3. Most of the estimates have confidence intervals that overlap with no effect (0% vaccine effectiveness) and even a negative effect (<0%). A negative effect suggests that vaccinated people are at increased risk of influenza infection compared to non-vaccinated people. Based on randomized clinical trials, vaccination is unlikely to cause increased risk of influenza infection, and the observation is therefore likely rather a result of uncertainty of the estimates, selection bias and confounding factors (being in a high-risk population influencing both vaccination status and risk of influenza infection).

**Figure 3.**
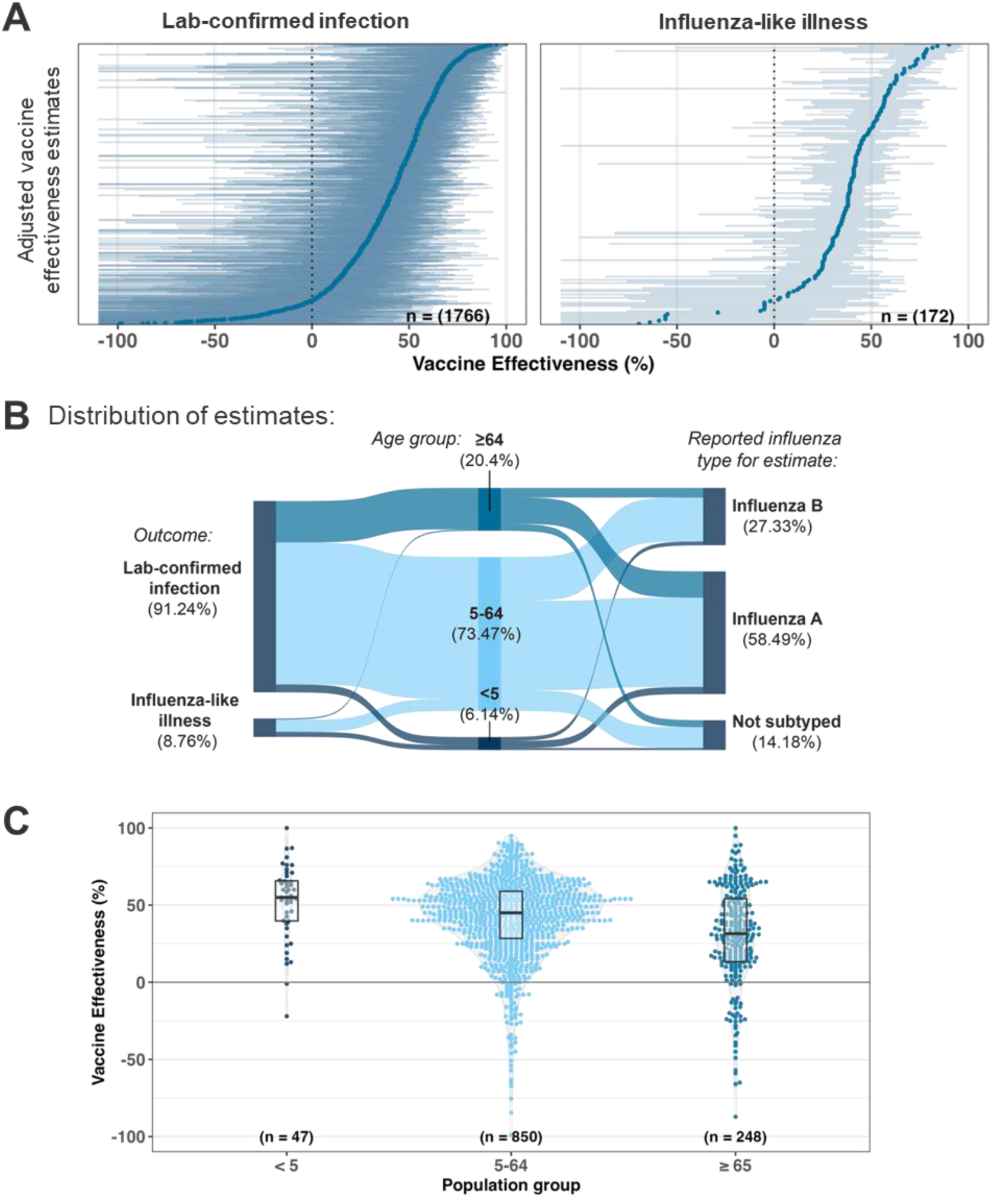
Influenza vaccine effectiveness estimates stratified by outcome, age group, and virus type. (A) Forest plots showing vaccine effectiveness estimates for outcomes defined by lab-confirmed infection (left) or influenza-like illness (right). Each estimate is shown as a dot, and the boundaries of the line indicate the lower and upper confidence intervals. (B) Relative proportion of estimates reported for influenza-like illness or lab-confirmed infection as an outcome, across different age groups, as well as different influenza virus types. (C) Distribution of vaccine effectiveness estimates for different risk groups defined by age (<5 years, 5-65 years, and >65 years of age).

### Meta-analysis

Vaccine effectiveness, the relative risk of influenza infection compared to non-vaccinated study participants, across all seasons and geographical locations was 39.4% overall (95% CI: 38.7–40.1%) and 41.2% for the general population (5-65 years; 95 % CI: 40.4–42.0%). It was higher in children under 5 years of age (51.7%; 95 % CI: 47.5–55.5%), but lower for people of 65 and older (27.4%; 95 % CI: 25.1–29.5%) (Table 1; Figure 3C).

**Table 1.** Vaccine effectiveness of seasonal influenza in risk groups defined by age and influenza virus. Lab-confirmed infection only. CI: confidence interval; (I)VE: (influenza) vaccine effectiveness. I^2^: level of heterogeneity (0% = no observed heterogeneity).

|  |  | All | Under 5's | General population | 65 and over |
| --- | --- | --- | --- | --- | --- |
| Influenza A | IVE (%) | 34.89 | 50.00 | 36.41 | 23.96 |
|  | Lower CI | 33.89 | 35.33 | 35.33 | 21.00 |
|  | Upper CI | 35.87 | 55.10 | 37.47 | 26.81 |
|  | Number of VE estimates | 685 | 27 | 496 | 151 |
|  | I<sup>2</sup> (%) | 66.93 | 60.54 | 68.89 | 51.60 |
|  | p | <0.001 | <0.001 | <0.001 | <0.001 |
| Influenza B | IVE (%) | 46.71 | 48.99 | 49.17 | 29.63 |
|  | Lower CI | 45.39 | 35.50 | 47.79 | 25.00 |
|  | Upper CI | 47.99 | 59.66 | 50.51 | 33.97 |
|  | Number of VE estimates | 316 | 12 | 242 | 56 |
|  | I<sup>2</sup> (%) | 53.72 | 50.04 | 49.04 | 31.81 |
|  | p | <0.001 | 0.024 | <0.001 | 0.014 |
| Any influenza | IVE (%) | 39.40 | 51.69 | 41.23 | 27.36 |
|  | Lower CI | 38.65 | 47.51 | 40.42 | 25.11 |
|  | Upper CI | 40.14 | 55.54 | 42.02 | 29.54 |
|  | Number of VE estimates | 1072 | 42 | 775 | 234 |
|  | I<sup>2</sup> (%) | 65.48 | 54.31 | 66.80 | 49.12 |
|  | p | <0.001 | <0.001 | <0.001 | <0.001 |

### Lower vaccine effectiveness against A/H3N2 compared to A/H1N1

The infecting influenza type (A or B) was available for a large subset of the studies (*N* = 120/145 articles, 82.8%). Vaccine effectiveness was higher against laboratory-confirmed influenza B (46.7%; 95% CI: 45.4%–48.0%) than influenza A (34.8%; 95% CI: 33.9%–35.8%) (Fig. 4A). In the group of ≥65 years, which had lower vaccine effectiveness overall, there was no significant difference for influenza A (24.0%, 95% CI: 21.0–26.8%) and B type (29.6%, 95% CI: 25.0–34.0%) (Table 1). Within influenza A infections, vaccine effectiveness was lower against A/H3N2 when compared to A/H1N1 across all populations (23.9%; 95 % CI: 22.3– 25.4%; vs 49.0%; 95 % CI: 47.8–50.2). Within influenza B, vaccine effectiveness against the Victoria lineage was lower when compared to that against the now extinct Yamagata lineage (43.6%, 95% CI: 39.9–47.2%vs 53.6%, 95% CI: 50.9–56.1%) across all populations (Table 2).

**Figure 4.**
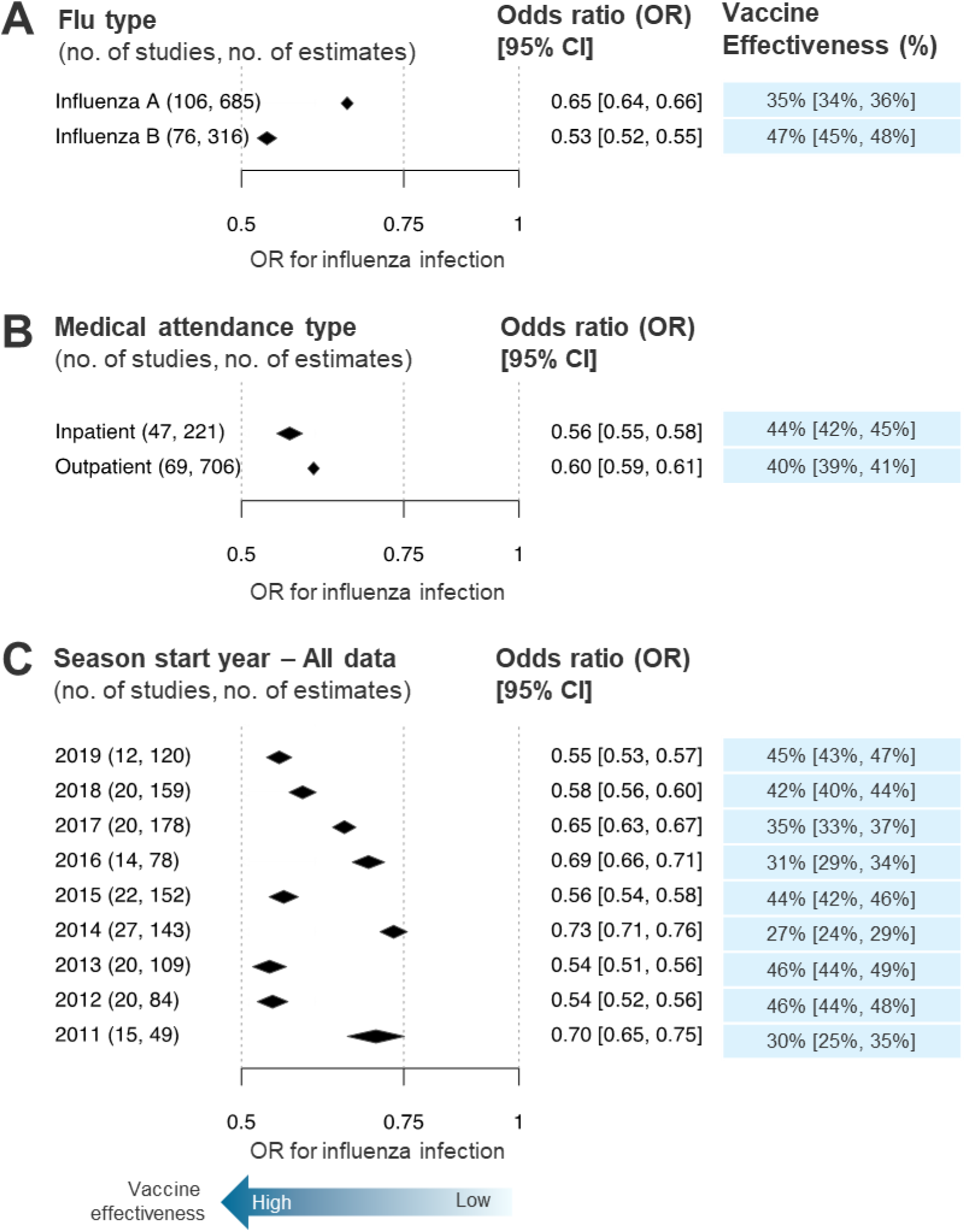
Forest plot summary representing the overall vaccine effectiveness of the meta-analysis. Each diamond indicates the relative odds of influenza infection for vaccinated compared to unvaccinated people (estimate and confidence interval) stratified by: (A) influenza A and B, (B) the inpatient and outpatient populations, and (C) influenza season start years between 2011 and 2019.

**Table 2.**
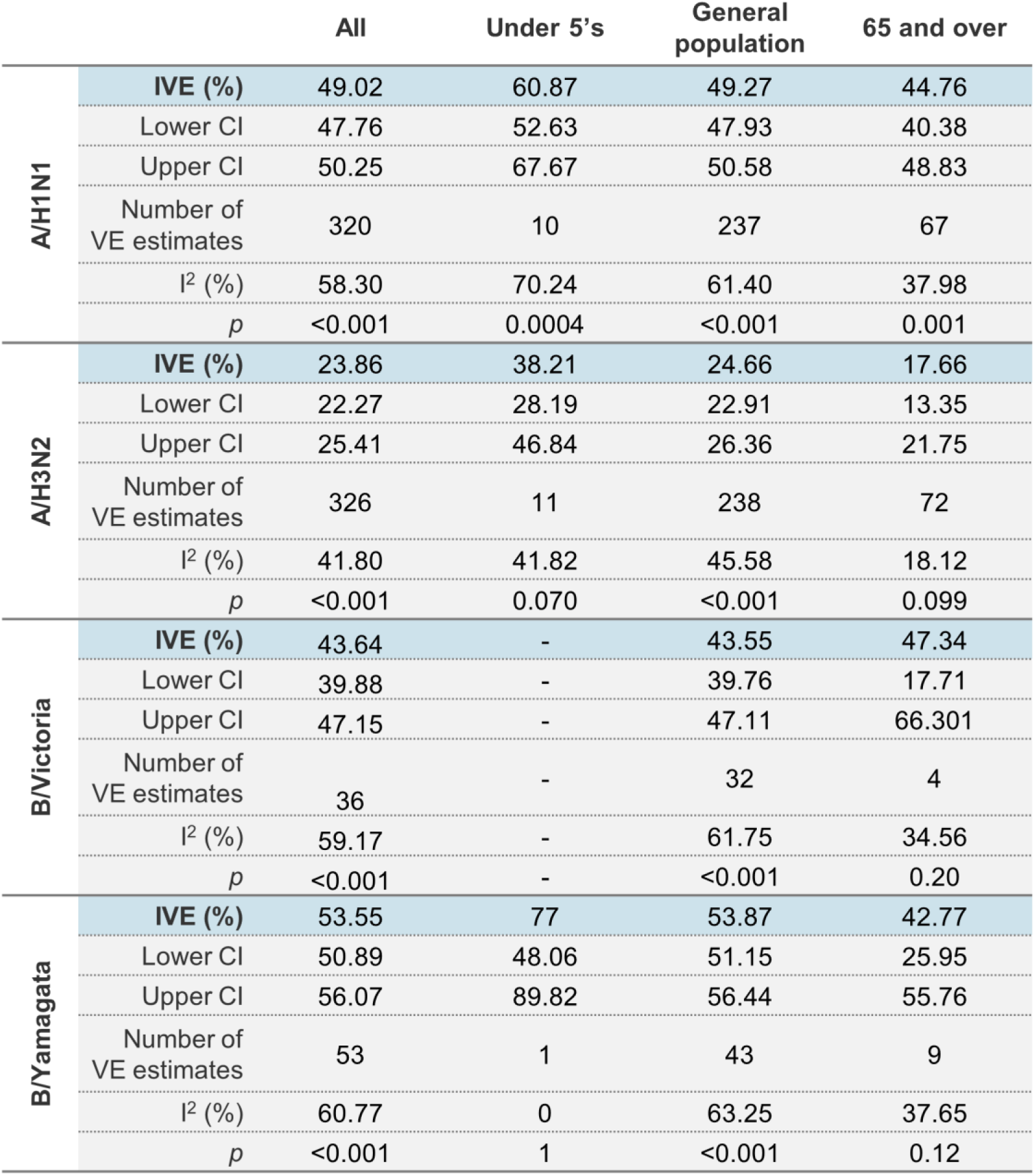
Vaccine effectiveness of seasonal influenza in risk groups defined by age and influenza subtype (as reported). CI: confidence interval; (I)VE: (influenza) vaccine effectiveness. I^2^: level of heterogeneity (0% = no observed heterogeneity).

### Lower vaccine effectiveness among outpatients compared to inpatients

When comparing medical attendance type (Fig. 4B), vaccine effectiveness was marginally higher amongst hospitalized patients (inpatients; 43.6%; 95 % CI: 41.7–45.1%) compared to outpatients (40.1%; 95% CI: 39.2–41.0%). For inpatients only, pooled vaccine effectiveness against influenza hospitalization was 43.3% (95 % CI: 40.8–45.8%) against influenza A, and 51.0% (95 % CI: 46.8–54.8%) against influenza B (Table 3). Focussing on participants from outpatient settings, pooled vaccine effectiveness was 34.8% (95 % CI: 33.6–35.9%) against influenza A, and 49.0% (95 % CI: 47.9–50.4%) against influenza B (Table 4).

**Table 3.** Vaccine effectiveness of seasonal influenza in risk groups defined by age and influenza virus. Inpatient groups only. CI: confidence interval; (I)VE: (influenza) vaccine effectiveness. I^2^: level of heterogeneity (0% = no observed heterogeneity).

|  |  | All | Under 5's | General population | 65 and over |
| --- | --- | --- | --- | --- | --- |
| Influenza A | IVE (%) | 43.33 | 78.76 | 44.84 | 37.32 |
|  | Lower CI | 40.78 | 56.72 | 41.98 | 31.41 |
|  | Upper CI | 45.78 | 89.58 | 47.57 | 42.71 |
|  | Number of VE estimates | 133 | 3 | 90 | 40 |
|  | I<sup>2</sup> (%) | 65.63 | 0.00 | 67.11 | 60.96 |
|  | p | <0.001 | 0.866 | <0.001 | <0.001 |
| Influenza B | IVE (%) | 50.98 | - | 52.24 | 41.18 |
|  | Lower CI | 46.83 | - | 47.90 | 25.96 |
|  | Upper CI | 54.82 | - | 56.22 | 53.27 |
|  | Number of VE estimates | 52 | - | 43 | 9 |
|  | I<sup>2</sup> (%) | 14.69 | - | 20.16 | 0.00 |
|  | p | 0.19 | - | 0.126 | 0.82 |
| Any influenza | IVE (%) | 43.59 | 57.66 | 45.50 | 38.63 |
|  | Lower CI | 41.74 | 46.60 | 43.31 | 34.62 |
|  | Upper CI | 45.38 | 66.43 | 47.60 | 42.39 |
|  | Number of VE estimates | 221 | 5 | 147 | 65 |
|  | I<sup>2</sup> (%) | 60.13 | 0 | 59.53 | 59.56 |
|  | p | <0.001 | 0.49 | <0.001 | <0.001 |

**Table 4.** Vaccine effectiveness of seasonal influenza in risk groups defined by age and influenza virus. Outpatient groups only. CI: confidence interval; (I)VE: (influenza) vaccine effectiveness. I^2^: level of heterogeneity (0% = no observed heterogeneity).

|  |  | All | Under 5's | General population | 65 and over |
| --- | --- | --- | --- | --- | --- |
| Influenza A | IVE (%) | 34.80 | 46.64 | 35.57 | 21.05 |
|  | Lower CI | 33.65 | 39.49 | 34.37 | 15.92 |
|  | Upper CI | 35.94 | 52.94 | 36.75 | 25.87 |
|  | Number of VE estimates | 464 | 16 | 364 | 82 |
|  | I<sup>2</sup> (%) | 67.20 | 73.25 | 69.38 | 33.14 |
|  | p | <0.001 | <0.001 | <0.001 | 0.003 |
| Influenza B | IVE (%) | 48.95 | 51.61 | 49.59 | 30.97 |
|  | Lower CI | 47.45 | 33.49 | 48.08 | 20.42 |
|  | Upper CI | 50.41 | 64.80 | 51.07 | 40.12 |
|  | Number of VE estimates | 220 | 7 | 181 | 31 |
|  | I<sup>2</sup> (%) | 52.49 | 67.58 | 52.95 | 28.11 |
|  | p | <0.001 | 0.005 | <0.001 | 0.075 |
| Any influenza | IVE (%) | 40.12 | 48.99 | 41.00 | 24.17 |
|  | Lower CI | 39.25 | 43.17 | 40.09 | 19.87 |
|  | Upper CI | 40.99 | 54.21 | 41.89 | 28.24 |
|  | Number of VE estimates | 706 | 23 | 559 | 121 |
|  | I<sup>2</sup> (%) | 66.87 | 71.12 | 69.12 | 25.18 |
|  | p | <0.001 | <0.001 | <0.001 | 0.008 |

### Variation between seasons and the impact of vaccine mismatch

Between seasons, there was marked variation in vaccine effectiveness. It ranged from 26.9% (95% CI: 24.3%–29.3%) in 2014 to a maximum of 46.4% (95 % CI: 44.0%–48.6%) in 2013 (Fig. 4C). The three years with the lowest vaccine effectiveness were 2011 (30.0%, 95% CI: 24.9%–34.9%), 2014 (26.9%, 95% CI: 24.3%–29.3%), and 2016 (31.3%, 95% CI: 28.5%–34.1%).

The degree to which vaccine strains match the circulating influenza strains significantly impacts vaccine effectiveness. Robust and standardized data on the circulating strains for each season is not available in many countries. In the United States of America (Fig. 5A), this information is provided annually by the CDC, specifically the levels of genetic and antigenic match between vaccine antigens and circulating strains, as summarized in Fig. 5B (Appendix III). Match is defined as the percentage of circulating influenza subtypes and/or lineages present in the seasonal vaccine component per year. Across all seasons, antigenic match was best for A/H1N1: always at least 95%. It was most variable for A/H3N2: ranging from 18.6% in 2014 to 100% in 2015. Vaccine effectiveness in the USA was lowest in 2014 (29.6%, 95% CI: 25.7%–33.2%), corresponding with the poor match in circulating A/H3N2 strains to the A/H3N2 vaccine component (18.6% match; Fig. 5B)

**Figure 5.**
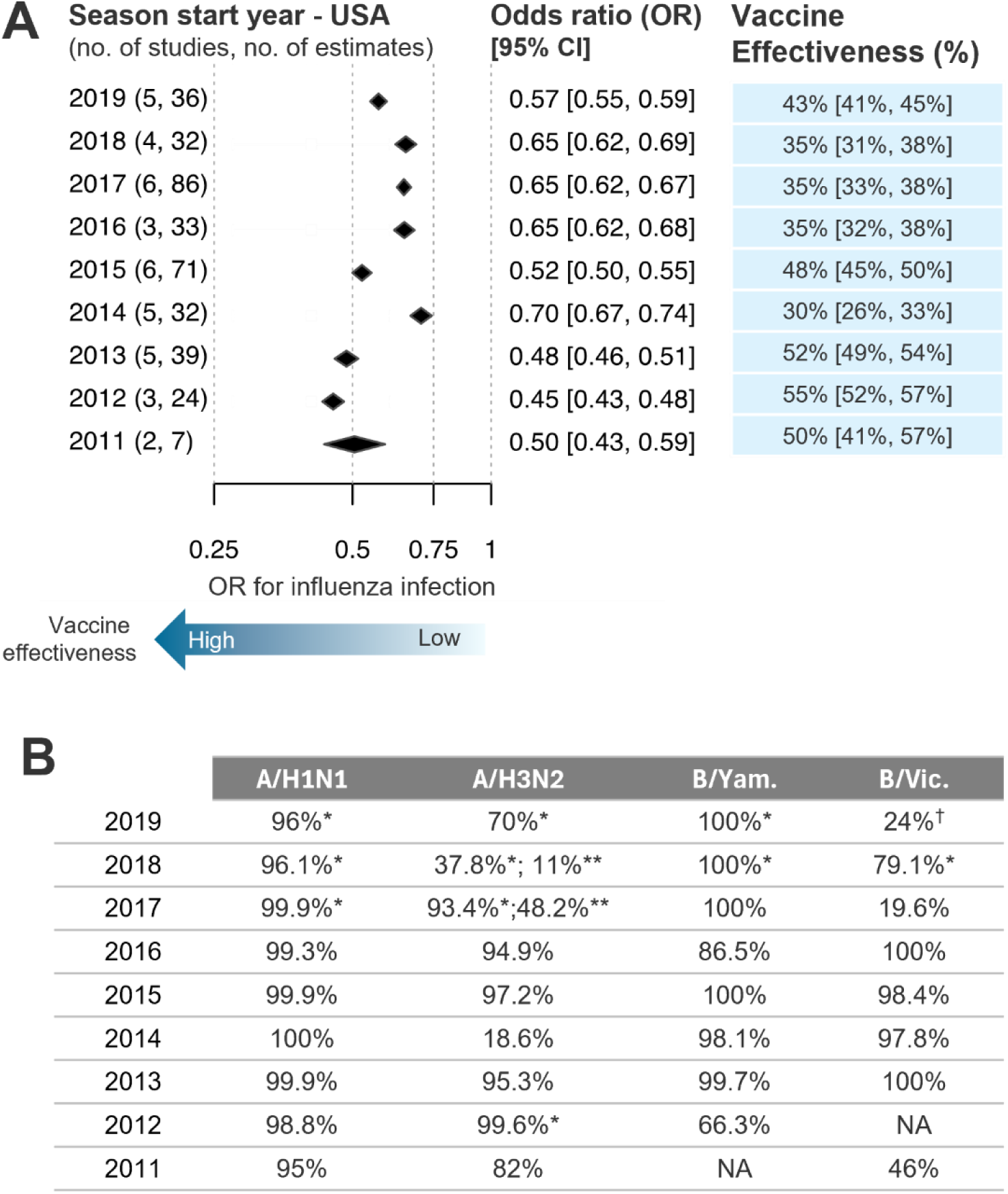
Overall vaccine effectiveness of the meta-analysis per season in the United States of America. (A) Each diamond indicates the relative odds of influenza infection for vaccinated compared to unvaccinated people (estimate and confidence interval) by influenza season start years between 2011 and 2019. (B) Table showing the percentage (%) of circulating influenza subtypes and/or lineages present in the seasonal vaccine component per year as reported by the CDC. Apart from 2019 B/Vic., which was genetically^†^ characterized, all other results were obtained from antigenic characterization. Viruses were propagated in cells* or eggs**. See Appendix III for the full excerpt of the CDC’s description of virus match or mismatch to the vaccine components across the influenza seasons. “100%” is used when the report states that “all” characterized viruses were similar to the vaccine virus component.

**Figure 6.**
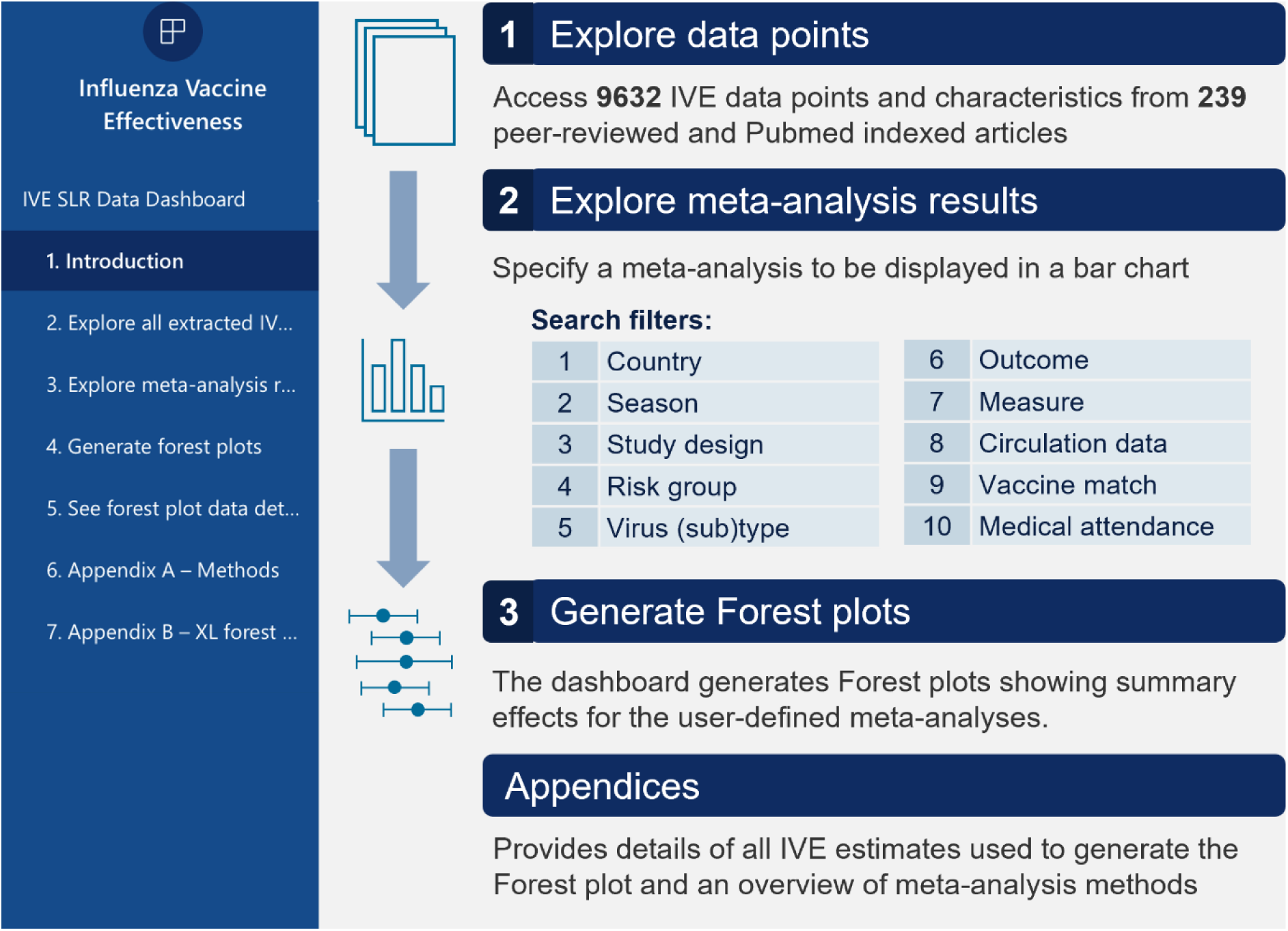
Schematic showing key functionalities of the dashboard. Screenshots of each feature of a shiny implementation of the dashboard are shown in figure S2.

## Discussion

We developed an open-source dataset for the meta-analysis of vaccine effectiveness of various types of influenza vaccines in different populations and settings. The dataset contains data for all influenza seasons from 2011 until 2019 to identify the efficacy gaps in vaccine-mediated protection in the absence of respiratory pandemics. Details on vaccine brand, type, and dose are reported for individual data points. The meta-analyses can be filtered by model, geographic region, influenza season, measure (adjusted vs unadjusted), age groups, comorbidity-based risk groups, influenza (sub)type or lineage, and population (age groups; inpatient versus outpatients). The dataset can be uploaded to a dashboard interface, for example, a R Shiny application. This may be useful for researchers in public health or in development of vaccines and antivirals.

We demonstrated the relevance of such a dashboard for addressing various research questions, by estimating pooled vaccine effectiveness for all studies using both fixed-effects and random-effects meta-analyses. Over time, the dataset could be extended to include more regions, provide more flexibility for selecting specific age groups, and include a methodological quality assessment. Ideally, the dataset should be updated yearly to include the latest available evidence on vaccine effectiveness.

Our analysis identifies various situations with suboptimal vaccine effectiveness. First, vaccine effectiveness was lowest in cohorts over 65 years of age. Previous studies also reported reduced vaccine effectiveness in older cohorts^8,16–18^, attributed to a deficiency in the differentiation and proliferation of vaccine-specific memory B cells after vaccination^19^.

Second, vaccine effectiveness is lower for A/H3N2, likely due to a higher frequency of antigenic drift ^20,21^.^21,22^. New antigenic variants of A/H3N2 viruses emerge every 3–5 years, whereas new variants of A/H1N1 and influenza B viruses appear less frequently, typically every 3–8 years^22^. The decreased efficacy against A/H3N2 could also be caused by antigenic sin in older individuals, who were not exposed to A/H3N2 during their childhood. In contrast, people born after 1968, when A/H3N2 emerged, are more likely to have been exposed to it in childhood^23^. Vaccine effectiveness against B/Yamagata, which was higher than that against influenza A, has become less relevant. Since the onset of the COVID-19 pandemic, no infections with B/Yamagata have been reported, and the WHO has now advised against including influenza B/Yamagata in vaccines for the upcoming Southern Hemisphere season^24^.

Third, in seasons with ‘mismatch’, antigenic difference between the circulating strains and those in the vaccine have lower vaccine effectiveness. The influenza season of 2014-2015 stood out with rates of outpatient illness, increased hospitalizations, and a notable percentage of deaths linked to lower respiratory tract infections^25^. The mismatch between distributed vaccines in the USA and circulating A/H3N2 strains was likely responsible for the low efficacy observed. Similarly, excess illness and deaths due to influenza in 2017-2018 was likely caused by the poor match between B/Victoria antigens in distributed vaccines and circulating B/Victoria strains. In addition to antigenic mismatch, B/Yamagata was also omitted from the inactivated trivalent influenza vaccine in a season where infections by this lineage outnumbered that by B/Victoria viruses^26^.

The difference in vaccine effectiveness in inpatient versus outpatient populations is more difficult to interpret. The higher vaccine effectiveness may be explained by hospitalized patients presenting with more severe disease, but there may also be a confounding effect. Previous observational studies have shown that the characteristics of patients presenting with acute respiratory illness in outpatient and inpatient settings can be markedly different, with hospitalized patients being comparatively younger or older and having a higher prevalence of comorbidities^27,28^. Furthermore, inpatient and outpatient settings may have different guidelines for influenza testing and surveillance. Longitudinal cohort studies might be better suited to such comparisons than the test-negative design studies included in our analysis.

In summary, our findings indicate that the influenza vaccine effectiveness is i) modest for younger age groups, and notably low for individuals over 65 years old; ii) lower against influenza A, especially A/H3N2, than influenza B; and iii) strongly reduced in years where the vaccine does not match the circulating strain. These results highlight an efficacy gap that should be complemented by additional healthcare intervention strategies. Universal vaccines^29^ that are under clinical development aim to provide broad-spectrum protection against seasonal influenza strains as well as pre-pandemic influenza viruses, both of which undergo continual antigenic evolution. However, to protect the elderly or other individuals with impaired vaccine-induced immunity, passive prophylaxis with broadly-neutralizing antibodies^30^ may confer more potent and fast-acting protection. Strategies to prioritize interventions against influenza based on their strengths in specific use cases should be developed, guided by evidence from large-scale empirical datasets.

## Data availability

The following data dashboard-ready files have been deposited on Github (https://github.com/LL-publications/FluVE-Datahub): a) Individual vaccine effectiveness estimates; b) Study descriptives of studies with test-negative design; c) and all meta-analyses. Please address any data inquires to the corresponding authors.

## Author contributions

AHJB and JG conceived the study. AHJB and DH developed the search strategy. DH and HB extracted data and conducted the meta-analysis. KLR, JCMM and ALB designed the data visualizations and conducted statistical analyses for those. AHJB, BJ and ALB supervised the study. DH, KLR, JF and AB drafted the first version of the article, with input from BJ. All authors reviewed the manuscript and approved it.

## Ethics statement

Ethical approval was not applicable for this study as it involved secondary analysis of published literature.

## Acknowledgements

We thank Jaco M. Klap and Alberto Garcia Hernandez for statistical input and validation of methodology.

