## Supplemental Materials for "Data dashboard to support meta-analyses on influenza vaccine effectiveness across regions, influenza seasons and outcomes"

### Supplementary Figures

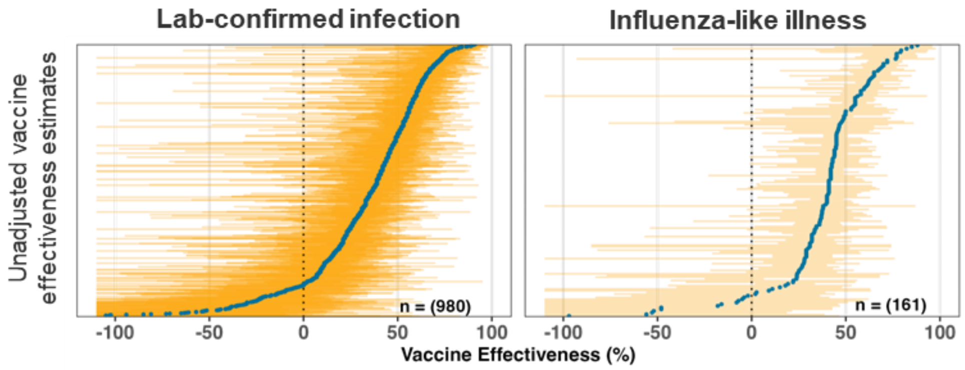


**Figure S1** Simplified forest plots showing unadjusted vaccine effectiveness estimates for outcomes defined by lab-confirmed infection (left panel) or influenza-like illness (right panel). Each estimate is shown as a dot, and the boundaries of the line indicate the lower and upper confidence intervals.

**Figure S2** Screenshots showing draft user interface and functionalities of an implementation of the dashboard.

### 1. Main page

- Describes the purpose and key functionality of the dashboard.

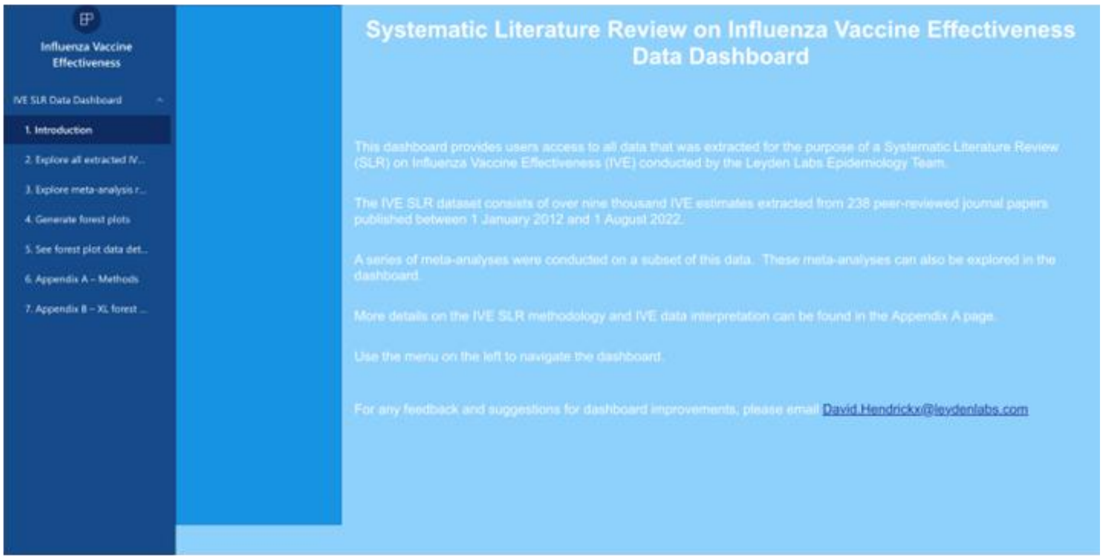


### 2. IVE estimates details page

- Provides access to all extracted IVE data points (n=9,632) and their characteristics.
- Each datapoint is linked to the relevant article's PubMed entry. The user can click the PubMed link to access the PubMed entry.
- The user can apply filters to query the dataset and find IVE data points of interest.

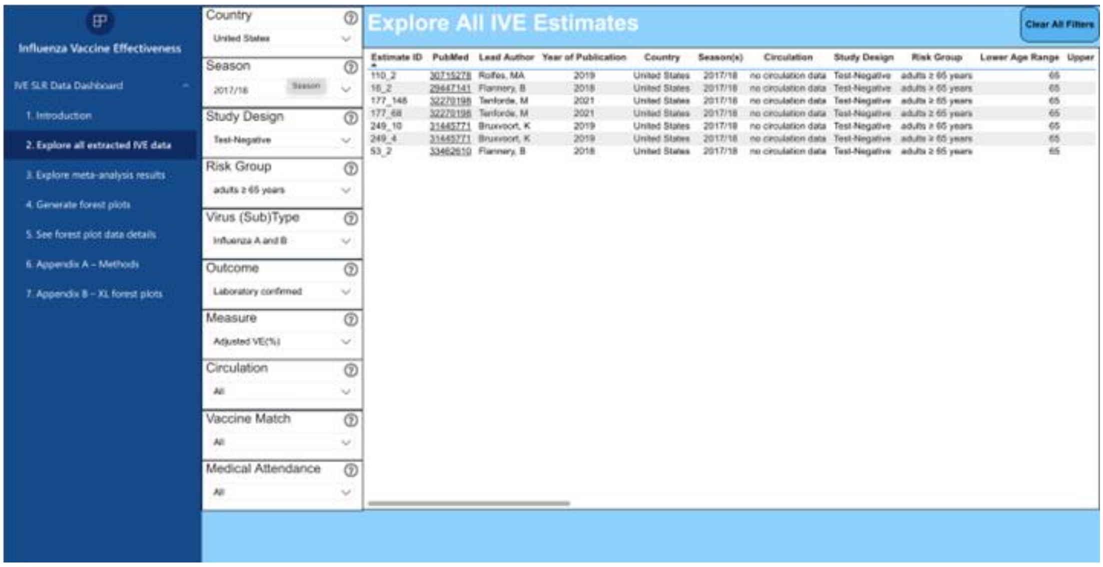

**3. Meta-analysis bar charts page**

- This page allows the user to specify a meta-analysis to be displayed in a bar-chart format.
- The meta-analyses are run for each influenza season for which there is data available.
- An aggregated 'all seasons' bar shows the overall meta-analysis result.
- Contextual mouseover popups summarise key data for each visualised meta-analysis, including the number of estimates included in each meta-analysis and the I2 statistic for heterogeneity.

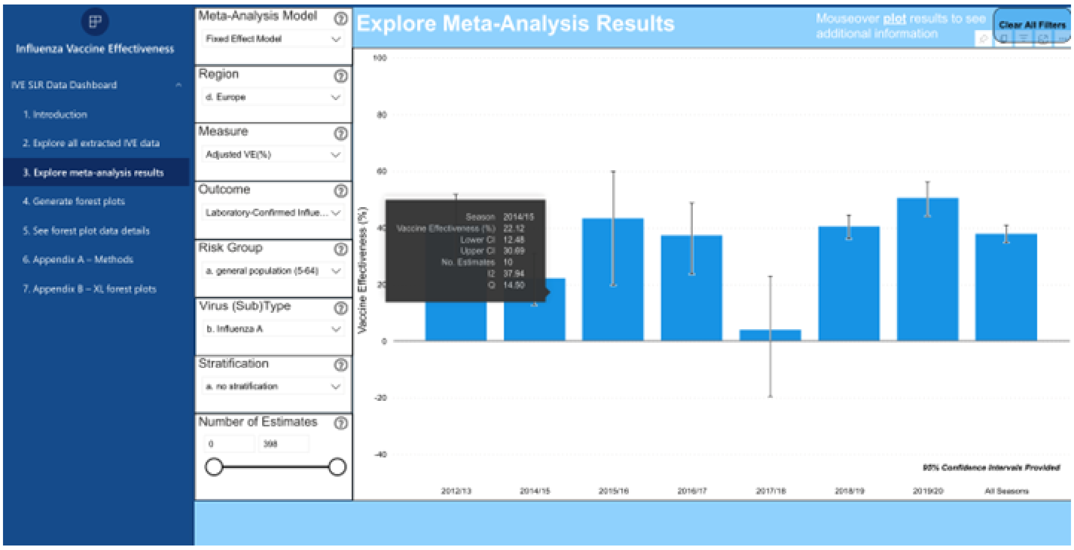


**4. Forest plots page**

- This page generates forest plots with corresponding summary effects for user-defined meta-analyses.
- Both fixed and random effect model forest plots are generated.

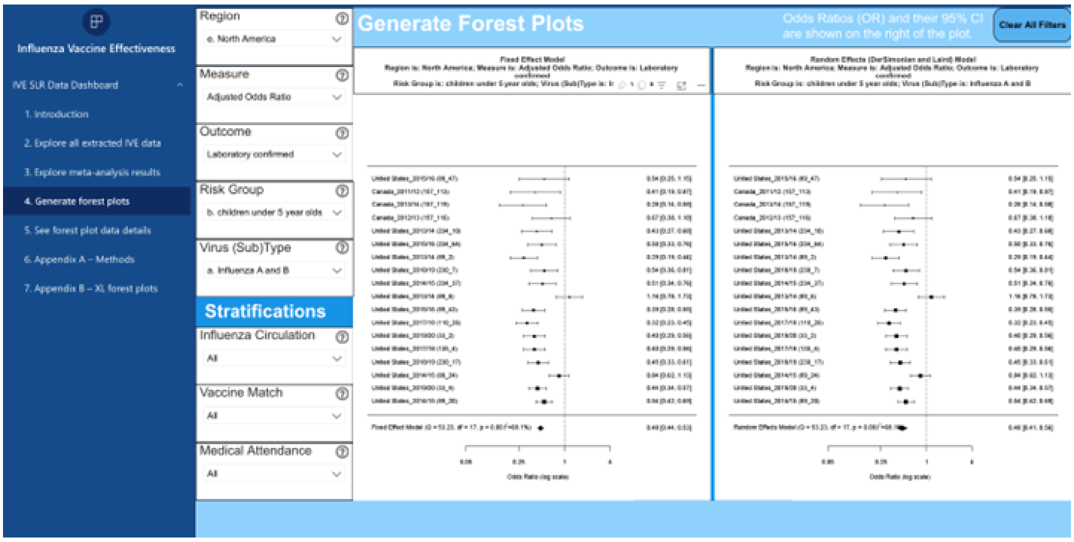

#### 5. Forest plots details page

- This page provides details on all IVE estimates that were included in the user-generated forest plot.

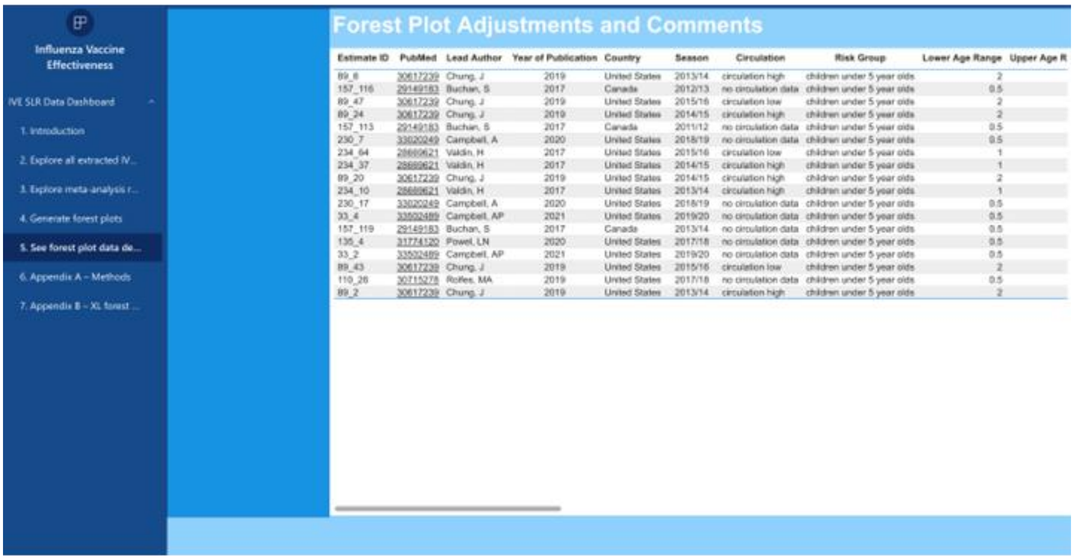


#### 6. Methods overview page

- This page provides an overview of the meta-analysis methods, including notes on data interpretation.

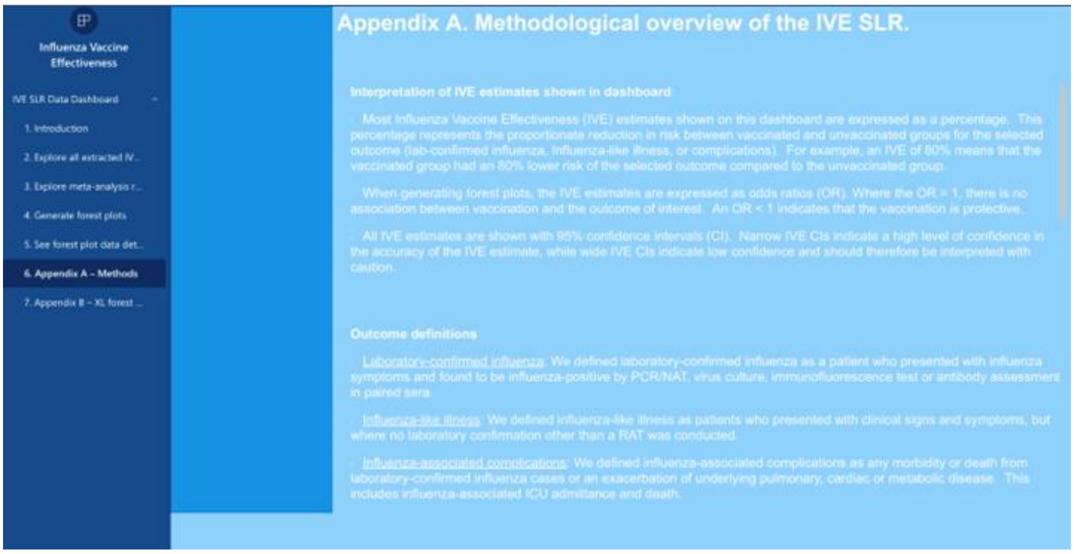

### Appendix I | Overview of methods

**1. Literature search parameters**

**2. Inclusion and exclusion criteria for studies included in the dashboard.**

**3. Inclusion and exclusion criteria for IVE estimates included in the meta-analyses.**

**4. Meta-analysis methodology and rationale**

**5. Outcome definitions used in the dashboard and meta-analysis**

### 1. Literature search parameters

**#0 Terms for defined search limits**

2012/01/01:2022/12/31[Date - Publication] AND ("humans"[MeSH Terms] AND ("dutch"[Language] OR "english"[Language] OR "french"[Language] OR "german"[Language] OR “spanish”[Language])) NOT "review"[Publication Type]

**#1 Terms for influenza**

Search String:

“Flu”[tiab] OR “Influenza-like illness”[tiab] OR “Influenza A virus”[Mesh] OR “Influenza B virus”[Mesh] OR “Influenza, Human”[Mesh]

**#2 Terms for vaccine effectiveness studies**

Search String:

“Vaccine Efficacy”[tiab] OR “Vaccine Effectiveness”[tiab] OR “Vaccine Efficacy”[Mesh]

**Final search:**

#0 AND #1 AND #2

### 2. Inclusion and exclusion criteria for studies included in the dashboard

The following **inclusion criteria** were applied:

- Reports data on influenza vaccine effectiveness outcomes of interest (laboratory-confirmed influenza or influenza-like illness)
- Conducted in European Economic Area countries, Switzerland, United Kingdom, United States, Canada, Australia, New-Zealand, Argentina, Chile, South Africa, Israel, Singapore, Hong Kong, Japan, South Korea or Taiwan
- Published in or after 2012
- Contains data collected on or after January 1, 2012
- Language: English, Dutch, French, German, Spanish

The following **exclusion criteria** were applied:

- Studies reporting on any of the following outcomes only: immunological parameters (e.g. antibody response); relative vaccine effectiveness across different influenza vaccines; unlicensed vaccines (including the trial of new adjuvants for licensed vaccines)
- Studies reporting on vaccination during pregnancy for the prevention of influenza in infants
- Non-human studies
- Case reports, case series, reviews and conference abstracts

### 3. Inclusion and exclusion criteria for IVE estimates included in the meta-analyses

| | Include | Exclude |
|---|---|---|
| **Study design** | • IVE estimates reported from studies using a test-negative study design. | • IVE estimates reported from studies using a design other than the test-negative design, such as cohort, generic case-control, and screening method study designs.<br>• IVE estimates that are stratified by data source and where the study also provides an overall IVE across all data sources. |
| **Outcomes** | • IVE estimates for any influenza (unspecified), influenza A, Influenza B, A/H1N1, A/H3N2, B/Victoria and B/Yamagata.<br>• IVE estimates for any of the pre-defined outcomes of interest: laboratory-confirmed influenza, influenza-like illness, and influenza-associated complications.<br>• IVE estimates for any medical attendance type, i.e., outpatient, inpatient and combined. | • IVE estimates for influenza subtype/lineage clades and subclades. |
| **Measure** | • Crude and adjusted IVE estimates expressed as a percentage, with their corresponding 95% confidence intervals.<br>• For adjusted IVE estimates where data is presented in a stepwise incremental manner with increasing number of adjustment covariates, include only the IVE estimate for the fully adjusted estimate. | • IVE estimates expressed in any measure other than percent, including odds ratios and relative risks.<br>• IVE estimates for which no 95% confidence intervals are provided.<br>• IVE estimates where non- numeric values have been used for the estimate or corresponding confidence intervals, e.g., *"< -25"* or *"-inf"*. |
| **Vaccine characteristics** | • IVE estimates for trivalent and quadrivalent seasonal influenza vaccines.<br>• IVE estimates for generic seasonal influenza vaccines where no further vaccine details are provided. | • IVE estimates that were stratified for the following vaccine characteristics: vaccine type (e.g., IIV vs LAIV); dosing; vaccine technology; use of adjuvants; strain make-up; vaccine administration route.<br>• In paediatric populations, exclude IVEs for partially vaccinated groups; include only IVEs for fully vaccinated groups (i.e., 2 doses). |
| **Risk groups** | • IVE estimates for all defined risk groups.<br>• IVE estimates for the general population (5 to 64 years) | • IVE estimates for generic 'other' risk groups<br>• IVE estimates for specific study populations, e.g., healthcare workers and military personnel.<br>• IVE estimates for defined risk groups, but which are further stratified by other characteristics, such as comorbidity presence, |

| | Include | Exclude |
|---|---|---|
| | | frailty, and time since last hospitalisation.<br>• IVE estimates for combined risk groups, such as children under 5 years who also have a cardiovascular condition. |
| **Patient history** | • IVE estimates for vaccination in the current reporting season, independent of vaccines received in previous seasons. | • IVE estimates that consider vaccination history prior to the reporting season, such as “vaccinated current season only” and “vaccinated previous and current season”.<br>• IVE estimates that are stratified by influenza treatment status (e.g., ‘Neuraminidase inhibitor treatment’).<br>• IVE estimates that are stratified by previous infection. |
| **Temporal factors** | • IVE estimates that are in reference to a single flu season. | • IVE estimates across two or more flu seasons.<br>• IVE estimates calculated for a period outside of the flu season, such as northern hemisphere summer.<br>• IVE estimates that are stratified by week, calendar month, or phase of the flu season.<br>• IVE estimates stratified by time between date of vaccination and date of swab, data of symptom onset or date of other outcome of interest (such as death).<br>• IVE estimates that are stratified by month of vaccination.<br>• Interim flu season IVE estimates when the reference (or a related reference) also reports on IVE estimates for the complete flu season. |
| **Demographic factors** | *na* | • IVE estimates that are sex specific.<br>• IVE estimates stratified by contextual family, sibling, and schooling characteristics.<br>• IVE estimates that stratify the main IVE results by race or ethnicity. |
| **Geographic factors** | • Country-specific IVE estimates for any of the pre-defined countries of interest. | • IVE estimates for two or more countries combined. |

### 4. Meta-analysis methodology and rationale:

- We used the Cochrane Handbook for Systematic Reviews of Interventions as our reference guide for conducting the meta-analysis.
- We followed the procedure detailed in section 6.3 of the Handbook, which recommends extracting estimates of effect directly (rather than extracting summary data) in certain situations. Several of these are relevant to our data:
    - Data is extracted from non-randomised studies.
    - Meta-analyses are conducted using adjusted effect estimates.
    - Summary data is not available for all effect estimates.
- Based on the above, we conducted the meta-analyses as follows:

1. As described above, we extracted the IVE values and their confidence intervals in % format. Our starting point is therefore each study's IVE estimate %, the corresponding lower CI %, and the corresponding upper CI %. All studies expressed 95% confidence intervals.
2. These three values expressed in % were transformed to their respective Odds Ratio. This was calculated as follows: OR = 1- (IVE% /100).
3. The inverse variance weighted method for meta-analysis requires log-transformed ORs to maintain symmetry. So for each study the OR value and corresponding CIs were log transformed prior to performing the ensuing steps.
4. The log-transformed CIs were used to calculate each study's standard error (SE). This was calculated as follows: SE = (Upper CI – Lower CI) / 3.92
   This formula for calculating the SE assumes 95% CIs and was taken from the Cochrane Manual, based on information provided in section 6.3.1 and 6.3.2.
5. The SE (still expressed in log transformed format) was used to calculate the variance (V). This was calculated as follows: V = SE^2 (i.e. SE squared)
6. The following two variables (in log transformed format) were used as inputs for the meta-analysis calculations:
    a. The log-transformed OR value
    b. The calculated variance V
7. The outcome of the meta-analysis calculations produces a summary effect in log-transformed OR format with 95% CIs. To transform this back to the original (and more intuitive) IVE % format, the following calculations were done:
    a. Take the exponential of the log-transformed OR and associated CIs.
    b. From this, calculate the IVE % as follows: IVE% = 1 – OR (and do the same for the CIs).

### 5. Outcome definitions used in the dashboard and meta-analysis

| Main reported outcomes | |
|---|---|
| Laboratory confirmed | • Concerns patients with clinical signs and symptoms confirmed as influenza infection by laboratory tests as specified below.<br>• INCLUDES: virus culture, PCR/NAT, immunofluorescence, antibody assessment in paired sera<br>• EXCLUDES: rapid antigen tests* |
| Influenza-like illness | • Patients who present with clinical signs and symptoms indicating ILI in the broadest sense, no laboratory confirmation was performed.<br>• INCLUDES: ILI patients with a positive influenza RAT only (and where there is no mention of PCR of culture). |
| Complications<br>*(this outcome is only included in the dashboard; it was excluded from our paper given limited availability of relevant IVE data)* | • Any morbidity and death from laboratory-confirmed influenza-related causes.<br>• INCLUDES: patients in whom the virus produces complicated illness (e.g., pneumonia, acute otitis media), or an exacerbation of underlying pulmonary, cardiac or metabolic diseases.<br>• INCLUDES: ICU admittance and death. |
| **Medical attendance type** | |
| Outpatient, laboratory confirmed influenza | • Any medical attendance type for laboratory confirmed influenza that is not a hospitalisation or ICU admittance. |
| Inpatient, laboratory confirmed influenza | • Any hospitalisation or ICU admittance for laboratory confirmed influenza. |
| Outpatient and inpatient, laboratory confirmed influenza | • Any medical attendance type for laboratory confirmed influenza where the study population consists both of out- and inpatients.<br>• INCLUDES: IVE estimates for study populations where attendance type is unspecified, but where it is likely that both out and inpatients were included based on contextual information provided. |
| na | • No details available on medical attendance type.<br>• Outcome is for deaths or other severe complications, such as pneumonia.<br>• Outcome is for non lab-confirmed influenza.<br>• Medical attendance is part of a clinical trial active surveillance method. |

* Unless when performed in a hospital setting and RATs are listed as only one of multiple laboratory tests, including PCR and/or culture.

### Appendix II | Influenza type recoding for meta-analysis

```
MAdata3 <- MAdata3 %>%
  mutate(est_virustype_rec2_all = case_when(
    str_count(stu_flutypes, ",") == 0 ~ "include",
    est_virustype == "h1n1" &
      ((!grepl("infa", stu_flutypes) & !grepl("all", stu_flutypes)) |
      (((grepl("infa", stu_flutypes) | grepl("all", stu_flutypes)) & grepl("h3n2", stu_flutypes) )))  ~ "include",
    est_virustype == "h3n2" &
      ((!grepl("infa", stu_flutypes) & !grepl("all", stu_flutypes)) |
      (((grepl("infa", stu_flutypes) | grepl("all", stu_flutypes)) & grepl("h1n1", stu_flutypes) )))  ~ "include",
    est_virustype == "byam" &
      ((!grepl("infb", stu_flutypes) & !grepl("all", stu_flutypes)) |
      (((grepl("infb", stu_flutypes) | grepl("all", stu_flutypes)) & grepl("bvic", stu_flutypes) )))  ~ "include",
    est_virustype == "bvic" &
      ((!grepl("infb", stu_flutypes) & !grepl("all", stu_flutypes)) |
      (((grepl("infb", stu_flutypes) | grepl("all", stu_flutypes)) & grepl("byam", stu_flutypes) )))  ~ "include",
    est_virustype == "all" &
      !grepl("h1n1", stu_flutypes) &
      !grepl("h3n2", stu_flutypes) &
      !grepl("bvic", stu_flutypes) &
      !grepl("byam", stu_flutypes) &
      !grepl("infa", stu_flutypes) &
      !grepl("infb", stu_flutypes) ~ "include",
    est_virustype == "all" &
      (
        ((!grepl("infa", stu_flutypes)) & (!grepl("h1n1", stu_flutypes) | !grepl("h3n2", stu_flutypes))) |
        ((!grepl("infb", stu_flutypes)) & (!grepl("bvic", stu_flutypes) | !grepl("byam", stu_flutypes)))
      ) ~ "include",
    est_virustype == "infa" & (!grepl("h1n1", stu_flutypes) | !grepl("h3n2", stu_flutypes)) ~ "include",
    est_virustype == "infb" & (!grepl("bvic", stu_flutypes) | !grepl("byam", stu_flutypes)) ~ "include",
    TRUE ~ "exclude"
  ))

MAdata3 <- MAdata3 %>%
  mutate(est_virustype_rec2_a = case_when(
    est_virustype == "h1n1" & (!grepl("infa", stu_flutypes) | (grepl("infa", stu_flutypes) & grepl("h3n2", stu_flutypes))) ~ "include",
    est_virustype == "h3n2" & (!grepl("infa", stu_flutypes) | (grepl("infa", stu_flutypes) & grepl("h1n1", stu_flutypes))) ~ "include",
    est_virustype == "infa" & (!grepl("h1n1", stu_flutypes) | !grepl("h3n2", stu_flutypes)) ~ "include",
```

```
    TRUE ~ "exclude"
  ))

MAdata3 <- MAdata3 %>%
  mutate(est_virustype_rec2_b = case_when(
    est_virustype == "byam" & (!grepl("infb", stu_flutypes) | (grepl("infb", stu_flutypes) & grepl("bvic", stu_flutypes))) ~ "include",
    est_virustype == "bvic" & (!grepl("infb", stu_flutypes) | (grepl("infb", stu_flutypes) & grepl("byam", stu_flutypes))) ~ "include",
    est_virustype == "infb" & (!grepl("bvic", stu_flutypes) | !grepl("byam", stu_flutypes)) ~ "include",
    TRUE ~ "exclude"
  ))
```

### Appendix III | US Vaccine matching summary by season

**2011/12**

**Trivalent vaccine; vaccine is missing a B/Yamagata component)**

From the annual MMWR influenza activity report:

- Since October 1, 2011, CDC has antigenically characterized 1,887 influenza viruses submitted by U.S. laboratories including 527 pH1N1 viruses, 1,058 influenza A (H3N2) viruses, and 302 influenza B viruses.
- **Of the 527 pH1N1 viruses tested, 503 (95%) were characterized as A/California/7/2009-like, the pH1N1 component of the 2011–12 influenza vaccine.** Twenty-four viruses (5%) of the 527 tested showed reduced titers with antiserum produced against A/California/7/2009.
- **Of the 1,058 influenza A (H3N2) viruses, 864 (82%) were characterized as A/Perth/16/2009-like, the influenza A (H3N2) component of the 2011–12 influenza vaccine for the Northern Hemisphere.** A total of 194 (18%) of the 1,058 tested showed reduced titers with antiserum produced against A/Perth/16/2009.
- **Of the 302 influenza B viruses tested, 147 (49%) belonged to the B/Victoria lineage, and 139 (95%) of these were characterized as B/Brisbane/60/2008-like, the influenza B component for the 2011–12 Northern Hemisphere influenza vaccine.** Eight (5%) of the 147 viruses belonging to the B/Victoria lineage showed reduced titers with antisera produced against B/Brisbane/60/2008. A total of 155 (51%) viruses tested belonged to the B/Yamagata lineage.

**2012/13**

**Trivalent vaccine; vaccine is missing a B/Victoria component**

From the MMWR influenza activity report:

- CDC has antigenically characterized 2,452 influenza viruses collected since October 1, 2012, and submitted by U.S. laboratories, including 252 pH1N1 viruses, 1,324 influenza A (H3N2) viruses, and 876 influenza B viruses.
- **Of the 252 pH1N1 viruses tested, 249 (98.8%) were characterized as A/California/7/2009-like, the influenza A(H1N1) component of the 2012–13 influenza vaccine**. Three viruses (1.2%) of the 252 tested showed reduced titers with ferret antiserum raised against A/California/7/2009.
- **Of the 1,324 influenza A (H3N2) viruses, 1,319 (99.6%) were antigenically similar to the cell-propagated A/Victoria/361/2011 reference virus; most viruses tested were cell-propagated. The H3N2 vaccine component for the 2012–13 Northern Hemisphere season was egg-propagated A/Victoria/361/2011;** the use of egg-propagated vaccine viruses is a current regulatory requirement for vaccine production. Five (0.4%) of the 1,324 tested showed reduced titers with antiserum produced against cell-propagated A/Victoria/361/2011.
- **Of the 876 influenza B viruses tested, 581 (66.3%) belonged to the B/Yamagata lineage, and were characterized as B/Wisconsin/1/2010-like, the influenza B component for the 2012–13 Northern Hemisphere influenza vaccine.** A total of 295 (33.7%) viruses tested belonged to the B/Victoria lineage.

**2013/14**

From the MMWR influenza activity report:

- CDC antigenically characterized 2,905 influenza viruses collected and submitted by U.S. laboratories since October 1, 2013, including 2,036 pH1N1 viruses, 426 influenza A (H3N2) viruses, and 443 influenza B viruses.
- **Of the 2,036 pH1N1 viruses tested, 2,033 (99.9%) were antigenically similar to A/California/7/2009, the influenza A (H1N1) component of the 2013–14 Northern Hemisphere influenza vaccines**. Three viruses (0.1%) of the 2,036 tested showed reduced titers with ferret antiserum raised against A/California/7/2009.
- **Of the 426 influenza A (H3N2) viruses tested, 406 (95.3%) were antigenically similar to A/Texas/50/2012, the influenza A (H3N2) component of the 2013–14 Northern Hemisphere vaccines.** Twenty (4.7%) of the 426 tested showed reduced titers with antiserum produced against A/Texas/50/2012.
- **Of the 443 influenza B viruses tested, 323 (72.9%) belonged to the B/Yamagata lineage, and 322 (99.7%) were antigenically similar to B/Massachusetts/2/2012, the influenza B component of the 2013–14 Northern Hemisphere trivalent and quadrivalent influenza vaccines.** One (0.3%) virus showed reduced titers with antiserum produced against B/Massachusetts/2/2012.
- **The remaining 120 (27.1%) influenza B viruses belonged to the B/Victoria lineage and were antigenically similar to B/Brisbane/60/2008, the influenza B component of the 2013–14 Northern Hemisphere quadrivalent influenza vaccine.**

**2014/15**

From the MMWR influenza activity report:

- WHO collaborating laboratories in the United States are requested to submit a subset of their influenza-positive respiratory specimens to CDC for further virus characterization. CDC has antigenically and/or genetically characterized** 2,193 influenza viruses collected and submitted by U.S. laboratories since October 1, 2014, including 59 influenza A (H1N1)pdm09 viruses, 1,324 influenza A (H3N2) viruses, and 810 influenza B viruses.
- **Of the 59 influenza A (H1N1)pdm09 viruses tested, all were antigenically similar to A/California/7/2009, the influenza A (H1N1) component of the 2014–15 Northern Hemisphere influenza vaccine.**
- **A total of 246 (18.6%) of the 1,324 H3N2 viruses tested have been characterized as A/Texas/50/2012-like, the influenza A (H3N2) component of the 2014–15 Northern Hemisphere influenza vaccine.**
  A total of 1,078 (81.4%) of the 1,324 viruses tested showed either reduced titers with antiserum produced against A/Texas/50/2012 or belonged to a genetic group that typically shows reduced titers to A/Texas/50/2012. The viruses that showed reduced titers to A/Texas/50/2012 belonged to multiple genetic groups; most but not all were antigenically similar to the influenza A (H3N2) virus selected in September 2014 for the 2015 Southern Hemisphere and in February 2015 for the 2015–16 Northern Hemisphere influenza vaccines, A/Switzerland/9715293/2013. A total of 948 of the 1,324 A (H3N2) viruses were further characterized; 889 (93.7%) were antigenically similar to A/Switzerland/9715293/2013, and fifty-nine (6.2%) showed reduced titers with antiserum produced against A/Switzerland/9715293/2013 virus.
  Of the 810 influenza B viruses tested, 582 (71.9%) belonged to the B/Yamagata lineage, and the remaining 228 (28.1%) influenza B viruses tested belonged to the B/Victoria/02/87 lineage.

- **A total of 571 (98.1%) of the 582 B/Yamagata-lineage viruses were characterized as B/Massachusetts/2/2012-like, which was included as an influenza B component of the 2014–15 Northern Hemisphere trivalent and quadrivalent influenza vaccines.** Eleven (1.9%) of the B/Yamagata-lineage viruses tested showed reduced titers to B/Massachusetts/2/2012.
  Among the 582 B/Yamagata lineage viruses characterized, 576 (98.9%) viruses were antigenically similar to B/Phuket/3073/2013 virus, the B/Yamagata lineage virus selected for the 2015 Southern Hemisphere influenza vaccine and 2015–16 Northern Hemisphere influenza vaccine. Six (1.0%) showed reduced titers with antiserum produced against B/Phuket/3073/2013 virus.
- **A total of 223 (97.8%) of the 228 B/Victoria-lineage viruses were characterized as B/Brisbane/60/2008-like, the virus that is included as an influenza B component of the 2014–15 Northern Hemisphere quadrivalent influenza vaccine.** Five (2.2%) of the B/Victoria-lineage viruses tested showed reduced titers to B/Brisbane/60/2008.

**2015/16**

From the MMWR influenza activity report:

- CDC has antigenically or genetically characterized 2,616 influenza viruses collected and submitted by U.S. laboratories since October 1, 2015, including 997 influenza A(H1N1)pdm09 viruses, 625 influenza A(H3N2) viruses, and 994 influenza B viruses.
- **Among the 997 influenza A(H1N1)pdm09 viruses characterized, 996 (99.9%) were found to be antigenically similar to A/California/7/2009, the reference virus representing the influenza A(H1N1) component of the 2015–16 Northern Hemisphere influenza vaccine.** One (0.1%) of the A(H1N1)pdm09 viruses tested showed a reduced titer to A/California/7/2009. Although all recent influenza A(H1N1)pdm09 viruses belong to hemagglutinin (HA) genetic group 6B, two genetic subgroups, 6B.1 and 6B.2, have emerged, with the majority of U.S. viruses belonging to 6B.1. To date, however, viruses from these genetic subgroups remain antigenically similar to the A/California/7/2009 virus component in the vaccine.
- **All 625 influenza A(H3N2) viruses were genetically sequenced, and all viruses belonged to genetic groups for which a majority of viruses antigenically characterized were similar to cell-propagated A/Switzerland/9715293/2013, the reference virus representing the influenza A(H3N2) component of the 2015–16 Northern Hemisphere vaccine. A subset of 318 influenza A(H3N2) viruses also was antigenically characterized; 309 of 318 (97.2%) were similar to A/Switzerland/9715293/2013.**
- **A total of 548 influenza B/Yamagata-lineage viruses were characterized, and all were found to be similar to B/Phuket/3073/2013, the reference virus representing the influenza B/Yamagata-lineage component of the 2015–16 Northern Hemisphere trivalent and quadrivalent vaccines.**
- **A total of 446 influenza B/Victoria-lineage viruses were characterized, and 439 (98.4%) were found to be similar to B/Brisbane/60/2008, the reference virus representing the influenza B/Victoria-lineage component of the 2015–16 Northern Hemisphere quadrivalent vaccine.** Seven (1.6%) of the B/Victoria-lineage viruses tested showed reduced titers to B/Brisbane/60/2008.

**2016/17**

From the MMWR influenza activity report:

- CDC has antigenically characterized 1,824 influenza viruses collected by U.S. laboratories since October 1, 2016 (296 influenza A(H1N1)pdm09, 772 influenza A(H3N2), and 756 influenza B viruses).
- **Among the 296 A(H1N1)pdm09 viruses, 294 (99.3%) were antigenically characterized as A/California/7/2009-like, the influenza A(H1N1)pdm09 component of the 2016–17 Northern Hemisphere vaccine**.
- **Among the influenza A(H3N2) viruses, 730 (94.9%) were antigenically characterized as A/Hong Kong/4801/2014-like, a genetic group 3C.2a virus recommended as the A(H3N2) component of the 2016–17 Northern Hemisphere vaccine**. Among 42 viruses that were antigenically different from A/Hong Kong/4801/2014-like viruses (i.e., reacted poorly with ferret antisera raised against reference viruses representing A/Hong Kong/4801/2014-like vaccine viruses), 36 (85.7%) belonged to genetic group 3C.3a, represented by the A/Switzerland/9715293/2013 reference virus, which was included as the A(H3N2) component of the 2015–16 Northern Hemisphere vaccine.
- Among influenza B viruses, **327 B/Victoria-lineage viruses were antigenically characterized using postinfection ferret antisera and among these, 283 (86.5%) were antigenically characterized as B/Brisbane/60/2008-like, a recommended influenza B component of the 2016–17 Northern hemisphere trivalent and quadrivalent influenza vaccines.** Among the 44 B/Victoria linage viruses that had reduced titers against B/Brisbane/60/2008-like viruses, 39 (88.6%) belong to the B/Victoria deletion variant subgroup.
- **All 429 (100%) B/Yamagata-lineage viruses tested were antigenically characterized as B/Phuket/3073/2013-like, the recommended influenza B component of the 2016–17 Northern Hemisphere quadrivalent influenza vaccines.**

**2017/18**

From the MMWR influenza activity report:

- CDC has genetically characterized 3,329 influenza viruses collected since October 1, 2017, including 832 influenza A(H1N1)pdm09 viruses, 1,313 influenza A(H3N2) viruses, and 1,184 influenza B viruses. A subset of these viruses was also antigenically characterized.
- Phylogenetic analysis of the hemagglutinin (HA) gene segments from 832 A(H1N1)pdm09 viruses collected since October 1, 2017, showed that all viruses belonged to subclade 6B.1 (Supplementary Figure 3, https://stacks.cdc.gov/view/cdc/54975). This has been the predominant HA clade in the United States since the 2015–16 season (3 ).
- **Of the 736 A(H1N1)pdm09 viruses analyzed using HI assays, 735 (99.9%) were well inhibited (i.e., reacted at titers that were within fourfold of the homologous virus titer) by ferret antisera raised against cell culture–propagated 6B.1 virus A/Michigan/45/2015, the reference virus representing the A(H1N1)pdm09 vaccine virus for the 2017–18 Northern Hemisphere influenza season.**
- A total of 1,313 influenza A(H3N2) viruses were sequenced, and phylogenetic analysis of the HA gene segments indicated that multiple clades/subclades were cocirculating (Supplementary Figure 3, https://stacks.cdc.gov/view/cdc/54975), with 3C.2a predominating. Viruses with the 3C.2a HA emerged at the end of the 2013–14 season and have remained the predominant clade since the 2014–15 season (4 ), undergoing continued genetic diversification each season. **Among 655 representative A(H3N2) viruses antigenically characterized by HI or FRA, 612 (93.4%) were well inhibited by ferret antisera raised against A/Michigan/15/2014 (3C.2a), a cell-propagated reference virus representing A/Hong Kong/4801/2014 (the A(H3N2) component of the 2017–18 Northern Hemisphere influenza vaccines)**. Only 6.6% of A(H3N2) viruses, the majority of which belonged to genetic clade 3C.3a, showed evidence of antigenic drift (i.e., had eightfold or greater reductions in HI or FRA titers compared with reference virus titers). **In contrast to the 93.4%**

**of A(H3N2) viruses that were well inhibited by ferret antisera raised against cell-propagated A/Michigan/15/2014, only 48.2% of viruses tested were well inhibited by ferret antiserum raised against the egg-propagated A/Hong Kong/4801/2014 reference virus representing the A(H3N2) vaccine component.** A higher proportion (77.3%) of viruses tested were well inhibited by ferret antisera raised against egg-propagated A/Singapore/INFIMH-16–0019/2016 reference virus, representing the A(H3N2) component recommended for the 2018 Southern Hemisphere and the 2018–19 Northern Hemisphere influenza vaccines.

- Phylogenetic analysis of 896 influenza B/Yamagata-lineage viruses showed that all HA gene segments belonged to clade Y3 (Supplementary Figure 3, https://stacks.cdc.gov/view/cdc/54975), which also predominated in the 2016–17 season (5 ). **All 824 B/Yamagata lineage viruses that were antigenically characterized were antigenically similar to cell culture–propagated B/Phuket/3073/2013, the reference virus representing the B/Yamagata-lineage component of quadrivalent vaccines for the 2017–18 Northern Hemisphere influenza season.**
- The HA gene segment of 288 influenza B/Victoria-lineage viruses sequenced and phylogenetically analyzed belonged to genetic clade V1A, the same genetic clade as the vaccine reference virus, B/Brisbane/60/2008. However, 234 (81.3%) viruses had a six-nucleotide deletion in the HA gene segment (encoding amino acids 162 and 163). Viruses like these, previously abbreviated as V1A-2Del and now designated as V1A.1, were first reported during the 2016–17 season (5 ). **Among 270 antigenically characterized influenza B/Victoria viruses, only 53 (19.6%) were antigenically similar to cell culture–propagated B/Brisbane/60/2008, the reference virus representing the B/Victoria lineage component of 2017–18 Northern Hemisphere vaccines.** All 217 B/Victoria viruses that were poorly inhibited by antisera raised to B/Brisbane/60/2008 (i.e., had eightfold or greater reductions in HI titers compared with reference virus titers) had the V1A.1 HA segment. Circulating B/Victoria lineage V1A.1 viruses were well inhibited by ferret antisera raised against B/Colorado/06/2017, a V1A.1 reference virus representing the influenza B component recommended for the 2018–19 Northern Hemisphere influenza vaccine.

**2018/19**

<u>From the MMWR influenza activity report:</u>

- CDC genetically characterized 2,750 influenza viruses collected and submitted** by U.S. laboratories since September 30, 2018, including 1,251 influenza A(H1N1)pdm09 viruses, 1,024 influenza A(H3N2) viruses, and 475 influenza B viruses. A subset of these viruses also was antigenically characterized.
- Phylogenetic analysis of the hemagglutinin (HA) gene segments from the 1,251 characterized A(H1N1)pdm09 viruses determined that all belonged to genetic subclade 6B.1A, which evolved from clade 6B.1.
  **Among 331 antigenically characterized A(H1N1)pdm09 viruses, 318 (96.1%) were well inhibited (reacting at titers that were within fourfold of the homologous virus titer) by ferret antisera raised against A/Michigan/45/2015 (6B.1), the cell culture–propagated reference virus representing the A(H1N1)pdm09 component for the 2018–19 Northern Hemisphere influenza vaccines.**
- Phylogenetic analysis of the HA gene segments of 1,204 sequenced influenza A(H3N2) viruses indicated cocirculation of multiple clades/subclades. Circulating viruses possessed HA gene segments that belonged to clade 3C.2a (66; 6.4%), subclade 3C.2a1 (201; 19.6%), or clade 3C.3a (757; 73.9%). The frequency of 3C.3a viruses increased from 12.7% of the A(H3N2) viruses collected and sequenced by November 2018 to 81.9% of those collected and sequenced during December 2018–May 2019.
  **Among the 505 A(H3N2) viruses antigenically characterized by focus reduction assays**

**with ferret antisera, 191 (37.8%) were well inhibited by ferret antisera raised against A/Singapore/INFIMH-16-0019/2016 (3C.2a1), a cell culture–propagated reference virus representing the A(H3N2) component of 2018–19 Northern Hemisphere influenza vaccines. However, only 43 (11%) of the 388 viruses tested were well inhibited by antiserum raised against egg-propagated A/Singapore/INFIMH-16-0019/2016 reference virus,** likely because of egg-adaptive amino acid changes in the HA protein of the egg-propagated virus. **Three hundred fourteen (62.2%) viruses were poorly inhibited by ferret antiserum raised against cell culture–propagated A/Singapore/INFIMH-16-0019/2016 reference virus** (at titers that were reduced eightfold or more when compared with the homologous virus); among those viruses, 312 (99.4%) belonged to clade 3C.3a, the prevalence of which increased throughout the season.

- Phylogenetic analysis of 203 influenza B/Yamagata lineage viruses determined that the HA gene segments belonged to clade Y3. **All 178 B/Yamagata lineage viruses antigenically characterized were well inhibited by ferret antiserum raised against cell culture–propagated B/Phuket/3073/2013, the reference virus representing the B/Yamagata lineage component of quadrivalent vaccines for the 2018–19 Northern Hemisphere influenza season.**
- Multiple genetically and antigenically distinct B/Victoria lineage viruses cocirculated during the 2018–19 season. Viruses with a two-amino acid deletion (162–163) in the HA protein belong to subclade V1A.1, and viruses with a three-amino acid deletion (162–164) in the HA protein belong to subclade V1A-3Del.
  Among the 272 influenza B/Victoria lineage viruses sequenced and phylogenetically analyzed, the HA gene segment belonged to genetic clade V1A (40; 14.7%), subclade V1A.1 (137; 50.4%), or subclade V1A-3Del (95; 34.9%).
  **Among 191 B/Victoria lineage viruses antigenically characterized, 147 (79.1%) were well inhibited by ferret antiserum raised against cell culture–propagated B/Colorado/06/2017-like V1A.1 reference virus representing the B/Victoria lineage component of the vaccines for the 2018–19 Northern Hemisphere influenza season.** Among the 44 (20.9%) viruses that reacted poorly, 17 were antigenically related to the previous vaccine virus B/Brisbane/60/2008 and belonged to clade V1A, and 27 belonged to subclade V1A-3Del.

**2019/20**

**Available reporting is limited.**

**Important note**: This being the Covid-19 influenza season, CDC flu reporting was impacted. The MMWR excerpt below was published in October 2019 and reports on the May-September 2019 period, suggesting a degree of mismatch between circulating and vaccine strains (particularly in the B/Victoria component). A 2021 paper on the 2019/20 season published by CDC authors (see here) also suggests that there was a vaccine mismatch this season, particularly in the H1N1 component in the 2nd half of the season.

From the MMWR influenza activity report:

- CDC genetically characterized 867 influenza viruses submitted by U.S. and international laboratories during May 19–September 28, 2019, including 263 influenza A(H1N1)pdm09 viruses, 427 influenza A(H3N2) viruses, and 177 influenza B viruses.
- All A(H1N1)pdm09 viruses belonged to genetic subclade 6B.1A. **Among 25 antigenically characterized A(H1N1)pdm09 viruses, 96% were similar** to the cell-culture propagated 2019–20 Northern Hemisphere vaccine virus component.**
- The 427 influenza A(H3N2) viruses analyzed belonged to either clades 3C.2a (354; 83%) or 3C.3a (73; 17%) (Figure 2). Multiple subclades within the 3C.2a clade cocirculated with the majority of viruses belonging to subclade 3C.2a1, with regional differences in which subgroup

of 3C.2a1 predominated. A(H3N2) viruses with a clade 3C.3a HA, which reemerged last season, continue to circulate in the WHO Region of the Americas. **Among the 74 representative A(H3N2) viruses antigenically characterized, 70% were similar to the cell-culture propagated 2019–20 Northern Hemisphere vaccine virus component.** Thus, although ferret antisera clearly distinguish antigenic differences between 3C.2a and 3C.3a viruses there is some cross-reactivity.

- All 21 of the influenza B/Yamagata lineage viruses analyzed belonged to clade Y3. **All seven B/Yamagata lineage viruses antigenically characterized were similar to the cell culture–propagated 2019–20 Northern Hemisphere vaccine virus component.**
- Multiple genetically and antigenically distinct B/Victoria lineage viruses cocirculated. Viruses with a two-amino acid deletion (162–163) in the HA protein belonged to subclade V1A.1, and viruses with a three-amino acid deletion (162–164) in the HA protein belonged to subclade V1A-3Del. Among the 156 influenza B/Victoria lineage viruses analyzed, the HA gene belonged to clade V1A (six viruses; 4%), subclade V1A.1 (37; 24%), or subclade V1A-3Del (113; 72%). **Among the 53 B/Victoria lineage viruses antigenically characterized, the V1A.1 viruses were similar to the cell culture–propagated 2019–20 Northern Hemisphere vaccine component. Ferret antisera raised to recent V1A.1 viruses, however, had reduced reactivity with many viruses expressing V1A and V1A-3Del HA proteins indicating some antigenic differences between viruses in the different B/Victoria lineage subclades**. Nevertheless, sera from humans vaccinated with a V1A.1 virus cross reacted well with V1A-3Del viruses.
